# Estimating Cumulative COVID-19 Infections by a Novel “Pandemic Rate Equation”

**DOI:** 10.1101/2020.08.17.20176602

**Authors:** David H. Hamilton

**Affiliations:** Mathematics, University of Maryland, College Park

## Abstract

A fundamental problem dealing with the Covid-19 pandemic has been to estimate the rate of infection, since so many cases are asymptomatic and contagious just for a few weeks. For example, in the US, estimate the proportion *P(t)* = *N/*330 where *N* is the US total who have ever been infected (in millions)at time *t* (months, *t* = 0 being March 20). This is important for decisions on social restrictions, and allocation of medical resources, etc. However, the demand for extensive testing has not produced good estimates. In the US, the CDC has used the blood supply to sample for anti-bodies. Anti-bodies do not tell the whole picture, according to the Karolinska Instituet [2], many post infection cases show T-cell immunity, but no anti-bodies. We introduce a method based on a difference-differential equation (dde) for *P(t)*. We emphasize that this is just for the present, with no prediction on how the pandemic will evolve. The dde uses only *x* = *x (s)*, which is the number/million testing positive, and *y* = *y (s)*, the number/million who have been tested for all time 0 *≤ s ≤ t* (months), with no assumptions on the dynamics of the pandemic. However, we need two parameters. First, *ρ*, the ratio of asymptomatic to symptomatic infected cases. Second, *τ*, the period of active infection when the virus can be detected. Both are random variables with distribution which can be estimated. For fixed *ρ*, we prove uniform bounds

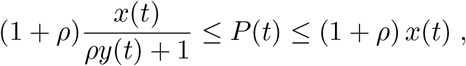

are best possible, with range depending on *τ*. One advantage of our theory is being able to estimate *P* for many regions and countries where *x* and *y* is the only information available.

## 1 Introduction

There has been some unfortunate modeling of the Covid epidemic[7]. We will stay away from predicting the future, but instead, try to understand the immediate past by using some biology, stats, and analysis, to develop a transparent formula to estimate the proportion *P (t)* of the US population that have *ever* been infected by the Coronavirus 19 at time *t* (months, where zero is March 20). This uses the number *y (s)* of those tested/million, and cases *x (s)*/million testing positive, over the time interval 0 *≤ s ≤ t*. The aim is to overcome the problem that testing misses those past the 1 *−* 3(?) week period of infection. The other problem is that asymptomatics are often not being tested, [14], [17], [18], [13]. Asymptomatics represent proportion *r* = *ρ/*(1 + *ρ)* of all infected, with the wide estimate 0.5 *< r* < 0.95. Ideally, one might try large scale testing for anti-bodies. However, many anti-bodies tests are unreliable, some infected do not develop antibodies. A recent study by King’s College [5] showed that a majority of post infection patients loose most of their anti-bodies within a month. In any case, there has been no widespread testing for anti-bodies. On June 24, the CDC [9] announced new estimates using sampling from regional US blood labs, where on average, 8% had anti-bodies. Considering the numbers testing positive in those regions, this implied *ρ ∼* 11. So, it is important to have a good estimate of *P*.

To reiterate, we introduce a new method to estimate *P (t)*, the proportion of the population who would test positive for the virus at any time before time *t*, i.e. all those who have ever been infected. This is a more robust measure as the pcr test, etc., for the virus is fairly reliable. Our method also requires parameter *τ*, the time period during which individuals test positive. We show *τ* is quite significant, indeed, we show how it affects the CDC estimate of *ρ*.

As “ for every infection only *ρ/*(1 + *ρ)* is symptomatic”, a simple estimate is

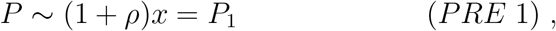

with PRE standing for “pandemic rate equation”. However, this assumes only symptomatic cases are counted, and ignores large scale testing which uncovers and counts asymptomatic cases.

Trying to count the asymptomatic cases, leads to the quadratic

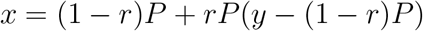

which has solution *P*_2_*(t)* =

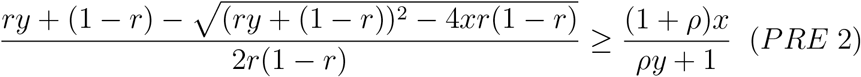

This second estimate assumes all testing done once at time *t*, whereas testing is spread over time. Furthermore, those who have been infected test positive only for a relatively short time, and testing past this period will not count them. Nevertheless, at the beginning of the pandemic, the bounds *P*_1_ and *P*_2_ were close. Now (at *t* = 4.6), we find for the US, assuming *r ∼*. 93 with *x*(4.6) = 1.5% and *y*(4.6) = 19.5%, that *P*_1_ *∼* 21.4% while *P*_2_ *∼* 6%.

Our main result models *P* as the solution of the “pandemic rate equation”:

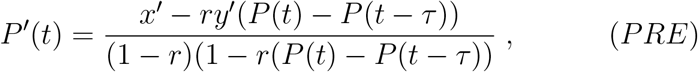

with initial conditions *x (t)* = *y (t)* = *P (t)* = 0*, t ≤* 0. Our assumption is that symptomatic cases are immediately treated, and thus tested (and counted). In actual fact, symptomatic cases take about a week before going for treatment/testing^2^. This delay could be entered as another random variable, but it is simpler to understand that our estimate will always be about a week out of date. Also, we assume that asymptomatic cases are discovered by essentially random testing from the entire population.

On June 24, the CDC[10] estimated *r* = .9125, i.e. the ratio of asymptomatic cases to symptomatic is about *ρ* = 11. The results of the Karolinska Inst.[2] shows *ρ ∼* 14 is more plausible, i.e. *r* = .93. Of course, *τ* and *r* are random variables whose distributions can be estimated. Simulations with lognormal distributed *τ* = 0.5 *±* 0.25*, r* = 0.93 *±* 0.03 (68% CI), our best estimate is

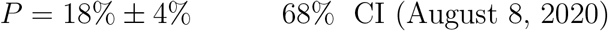

with the main error coming from uncertainty in *ρ*.

## 2 The Theory

Now for the mathematics, first the complete statements, then the proof.

### 2.1 The Formulae

As before, *P* = *P (t)* is the proportion of those in the population who would ever test positive for the virus in time interval [0*, t*] months, with initial time *t* = 0, March 20. As before, the parameters *r* and *τ* denote the proportion of the infected who are asymptomatic, and the length of infection, respectively. To begin with, we now assume the parameters *r*and *τ* are fixed for the population. The functions *x, y*, are smoothed with derivatives *x*′ *y*′.

We prove that *P* satisfies a nonlinear difference-differential equation:

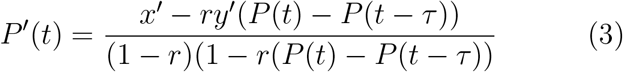

with initial conditions *x (t)* = *y (t)* = *P (t)* = 0*, t ≤* 0. The two simple minded estimates are essential bounds for the general case.

#### THEOREM

*Suppose that x, y, P, are increasing functions. Then, for fixed ρ, independently of τ the solution of (3) satisfies the bounds*

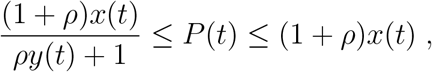

*and, furthermore, these bounds are best possible*.

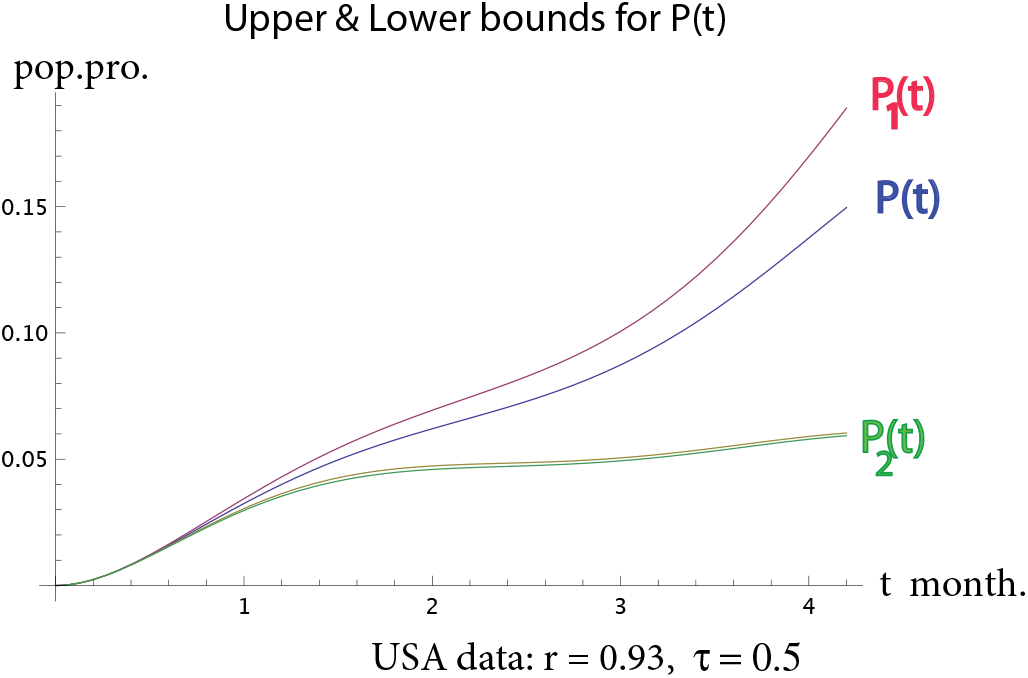

### 2.2 Derivation of Pandemic Rate Equation

We prove PRE. There is a large population of total size *n* million. Also, we use *X* = *X (s)* as the number who have tested positive, and *Y* = *Y (t)* as the number who have been tested up till time *s ≤ t* (months). As before, the parameters *r, τ*, denote the proportion of the infected who are asymptomatic, and the length of infection, respectively. We now assume the parameters *r, τ*, are fixed for the population of size *n*. People are sick for a time interval [*t − τ, t*] when they test positive. So, it is only during this time interval that the virus can be detected. Consider a very short time interval [*t −* ∆*t, t*], during which the quantities *x, y* change by ∆*x* = *x (t) − x (t −* ∆*t)*, ∆*y* = *y (t) − y (t −* ∆*t)*. The data for testing *y* and cases *x* is discrete and discontinuous, but we assume to be differentiable. Thus ∆*x ∼ x^0^ (t)*∆*t*, ∆*y ∼ y^0^ (t)*∆*t*.

The total number of cases comes from those who are symptomatic, take themselves for treatment and hence are tested; and those, who we assume are randomly tested, and a certain proportion turn out to be infected (but asymptomatic). We assume symptomatic and asymptomatic are only infected for a time period of length *τ* during which they test positive. There is the question of how quickly symptomatic cases go for treatment. We assume they immediately go for testing.^3^

We also use *Z (t)* the total number of symptomatic cases, and *W (t)* the number of asymptomatic cases. Evidently, *X* = *Z* + *W*. At time *t*, the number of past infections is *nP (t)*, so in time period [*t −* ∆*, t*], the number of new symptomatics is (1 *− r) nP^0^ (t)*∆*t ≥* 0, assuming *P* increasing. Thus,

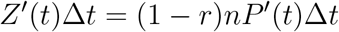

There is also ∆*Y ∼ Y^0^ (t)*∆*t* new tests, which counts those who sought help, and those asymptomatic cases caught up in essentially random sampling. Thus, not counting the symptomatics gives

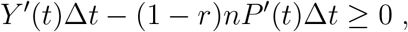

assuming *Y*′ ≥ *Z*′. The proportion of these testing positive is

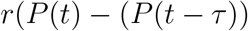

Thus, the new asymptomatics is

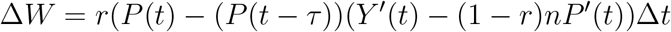

Now, as ∆*X* = ∆*Z* +∆*W*, the new cases during [*t−*∆*, t*] is ∆*X ∼ X^0^ (t)*∆*t* =

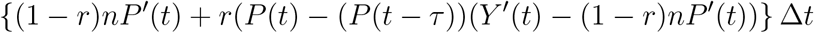

With ∆*t →* 0, and *x* = *X/n, y* = *Y/n* gives the delay-differential equation

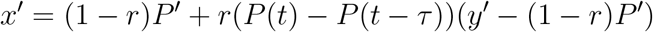

which simplifies to (3).

The nonlinear dde cannot be solved explicity in general, but is well suited for numerical solutions such as NDSolve in Mathematica, which has routines for delay-differential equations. The formula is for fixed *ρ* and *τ*, but in reality there is a distribution of values. We handled these by stochastic simulations.

### 2.3 Proof of THEOREM

This was motivated by considering two extreme cases. First case: *τ* = 0 *⇒ P (t) − P (t − τ)* = 0, and the dde becomes

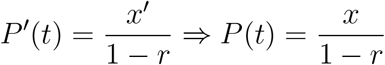

Secondly: once infected always infected, i.e. *τ* = *∞ ⇒ P (t − τ)* = 0, dde is

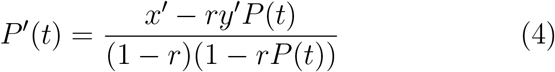

We are assuming *x, y*, are both are increasing with *x*′ ≤ *y*′. Writing *y* = *x*+*u* we have *u′>* 0 and 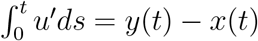 viewed as constraints on *u*. Hence,

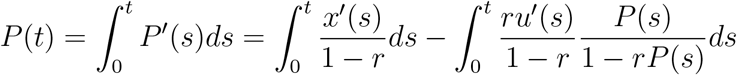

Now 0 *< P (s)* < 1 is increasing so

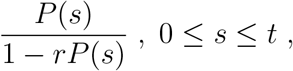

is maximized at *s* = *t*. Therefore, the second integral is maximised if *u*′ is a Dirac measure concentrated at *s* = *t*. It follows that *P* has lower bound

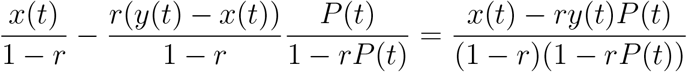

Solving for *P (t)* gives

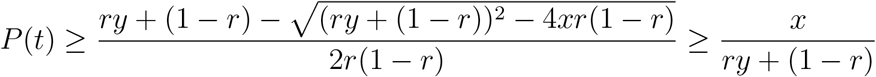

which proves the lower bound. Observe that for smooth *x, y*, this lower bound is not achieved, even in the limit as *τ → ∞*. Indeed, numerical solution of (4) for given *x* and *y* provides a somewhat better lower bound than *P*_2_.

Finally, we prove *P (t)* is sandwiched between *P*_1_ and *P*_2_ by showing the solution is monotone decreasing in *τ*. Consider

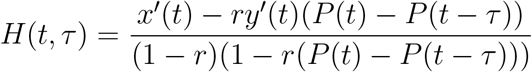

Now, as 0 *< r* < 1 and *x*′ < *y*′ we see that *H ≥* 0 as 0 *< P (t) − P (t − τ*) < 1. As *τ* varies from 0 to *∞*, we observe *P (t) −P (t−τ)* increases from 0 to *P (t)*. Define function

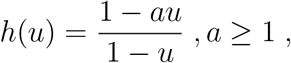

which is decreasing as

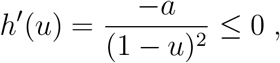

Thus *P*′ = *H* is monotone decreasing in *τ*. Hence, *P* decreases as *τ* increases.

We also carried out simulations for varying *τ* which gives consistent results:

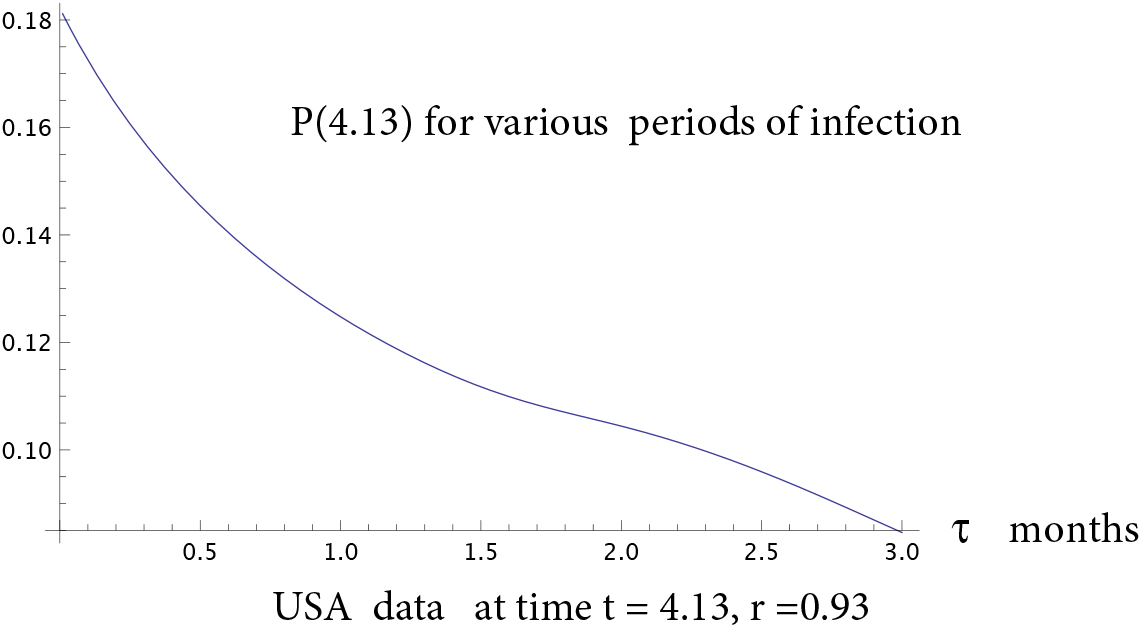

### 2.4 Estimating *ρ* and *τ*

The estimate of *ρ* (or *r)* is from a variety of sources, collected in [12]. In China, for children *r ∼*. 95; at a NYC hospital where all pregnant women were tested *r ∼*. 9; while *r ∼*. 5 for passengers on cruise ships. WHO currently has *r ∼*. 8 which is consistent with results from Iceland, where it is estimated *r >* 0.8 (where many “symptomatics” really had the flu which reduced the ratio).

We expect *r* to be different for different populations, indeed, be age dependent. Nevertheless, we want an average value for the entire population. By our formula, for early in the epidemic, we have *P ∼* (1 + *ρ) x*. Now, on March 29, for NYC, the CDC[9], [8] used anti-bodies to estimate *P ∼*. 07, while *x ∼*. 004, giving *r ∼*. 943. Later, on May 29, the CDC[10] estimate was *P ∼*. 23, while *x ∼*. 024, giving *r ∼*. 9. Similar results were found in different regions of the US giving the CDC average estimate *r* = 0.9. However, the Karolinska Inst. [2] estimates at least 25% more infections from their study of T-cell immunity. So, we take *r* = 0.93 *±* 0.03.

We make the important observation that *P ∼* (1 + *ρ) x* is only valid for small *x, y, t*, so using it to estimate *ρ* on present US data underestimates *ρ* by *∼* 20%. This underestimate will grow as *x* and *y* become larger.

## 3 Results

Our method was first tested on a hypothetical case. Then we use USA data.

### 3.1 Test cases

As nobody knows the true level of infection, we test by using a hypothetical infection which has proportion *P*_0_ in the population governed by:

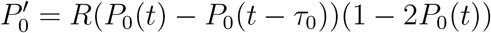

This assumes about 50% of the population can be infected (exactly once) and the period of contagion is *τ*_0_. *R* is the rate of infection. Also some small initial value *P*_0_*(t)* = *φ (t), t* < 0 is required. For *R* = 5 and *τ* = 0.25, this was solved numerically:

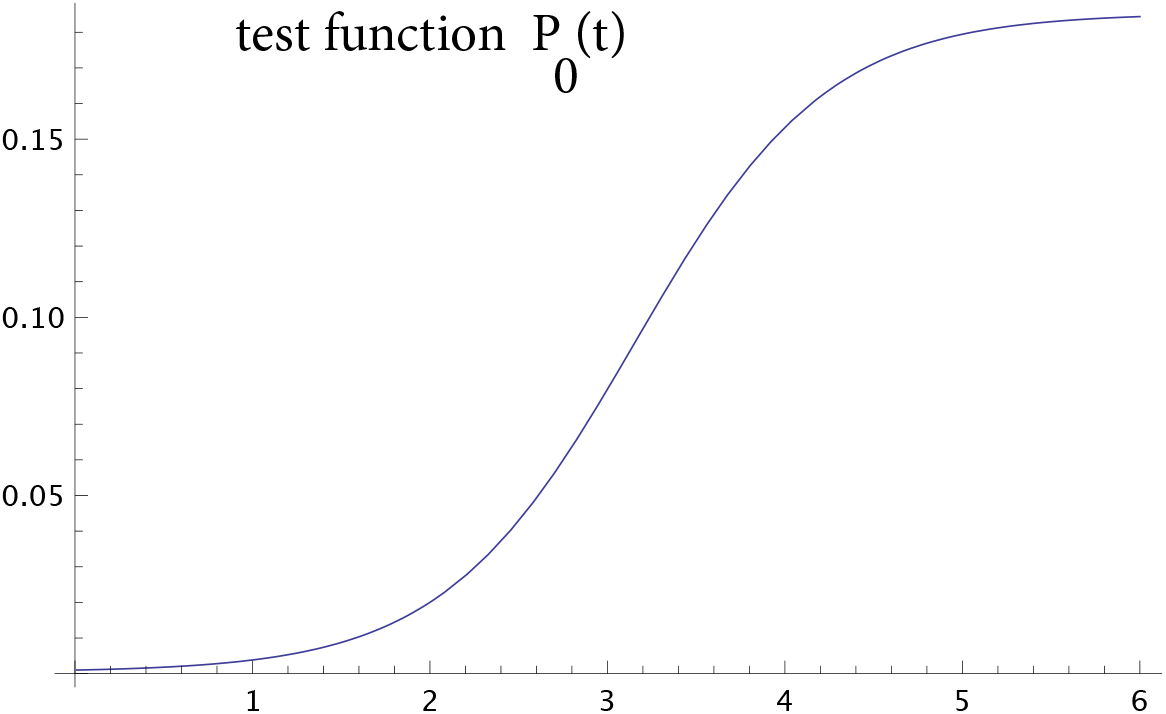

We need some testing function *y′ (t)*, choose *y*′ = 0.1 constant. Then, the function *x* for the number of cases/mill. satisfies the dde

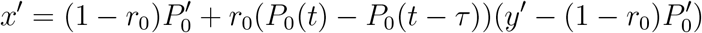

where the proportion *r*_0_ = 0.9 or *ρ*_0_ = 9 is given.

Next, we apply our process: first approximate *x*′, *y*′ by *X*1*, Y* 1. A not very good approximation is shown:

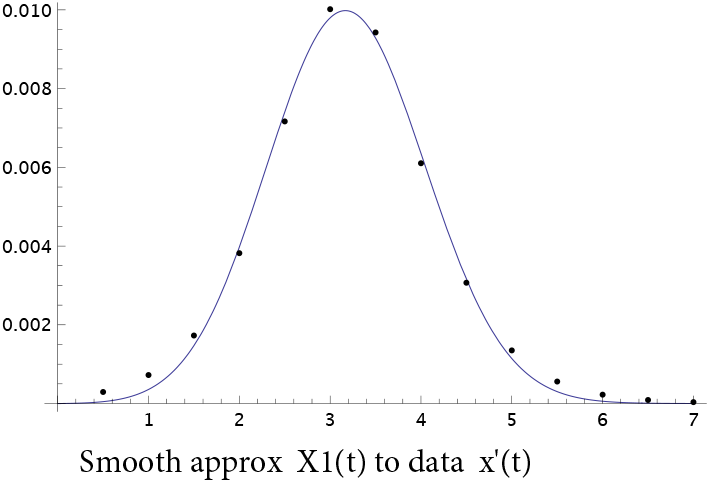

Then, obtain the predicted proportion *P (t)* using PRE:

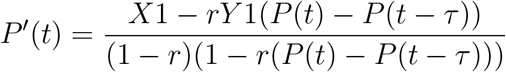

Now, of course, we have no apriori way of choosing the right *ρ* and *τ*. So these “would be obtained by field data”. However, if we choose the right ones, in this case *τ* = 0.25*, ρ* = 9

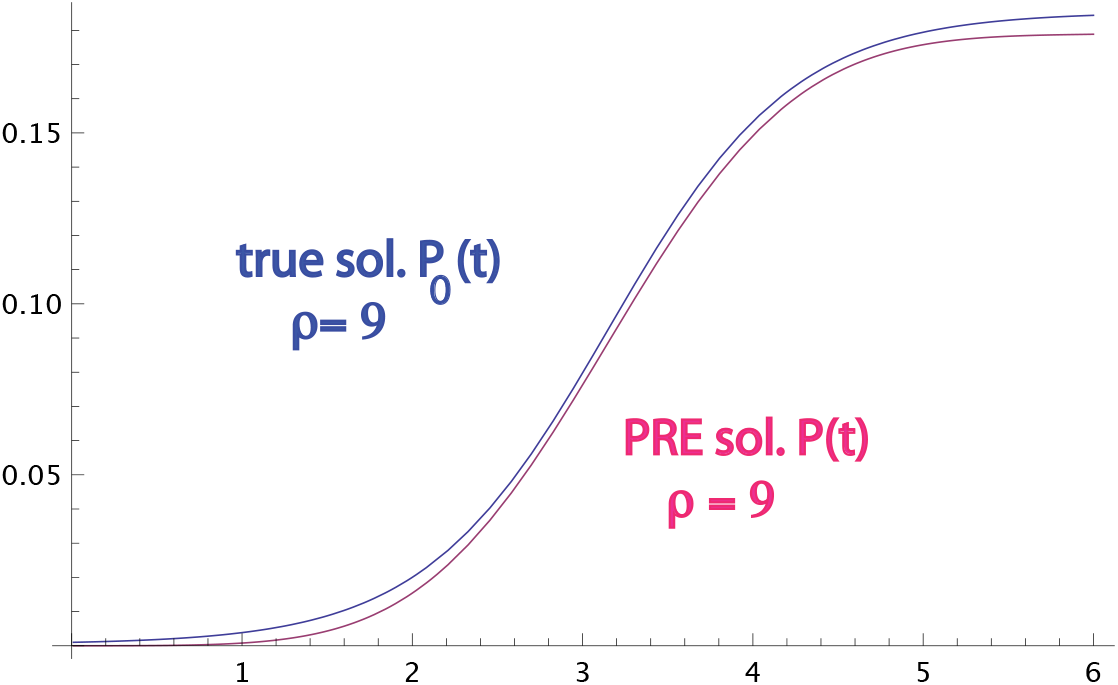

We find if *r, τ*, are close to the true *r*_0_*, τ*_0_, then the predicted proportion *P* is close to the true proportion *P*_0_. However, if say *r* is twice *r*_0_, then *P* is approximately twice the value of *P*_0_. On the other hand, the solution *P* is relatively less sensitive to variations in the period of infection *τ*.

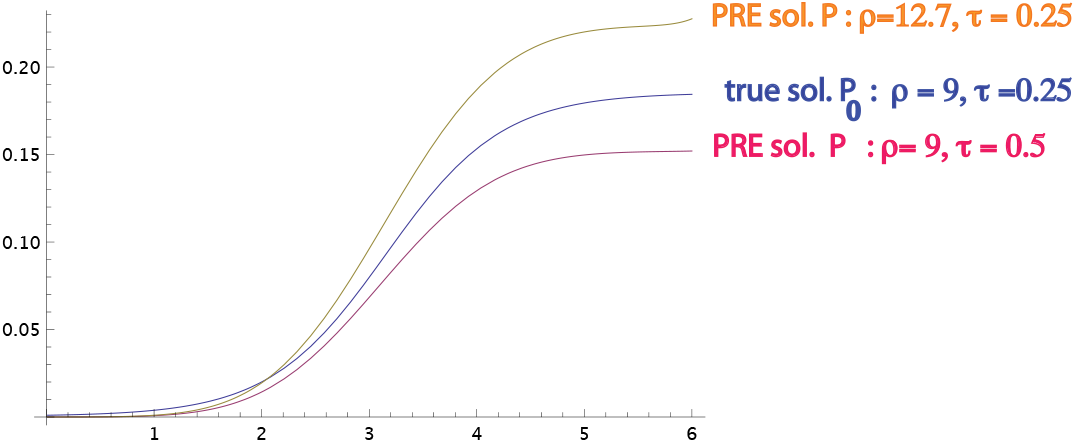

What this means is that the PRE is well posed, indeed, it is linear in *x*′, *y*′ and roughly linear in *ρ*.

### 3.2 USA data

For the US over time, 0 *≤ t ≤* 4.6 months, the data is approximated by

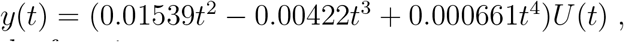

and by the function

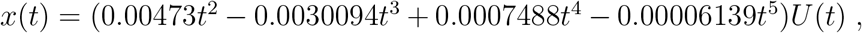

using the Heaviside Step Function *U (t)* to emphasize *x, y* = 0 for *t ≤* 0. This is simply a smooth interpolation and not meant to be any prediction, although the 3-4 term polynomials are a good fit for data at 11 points.

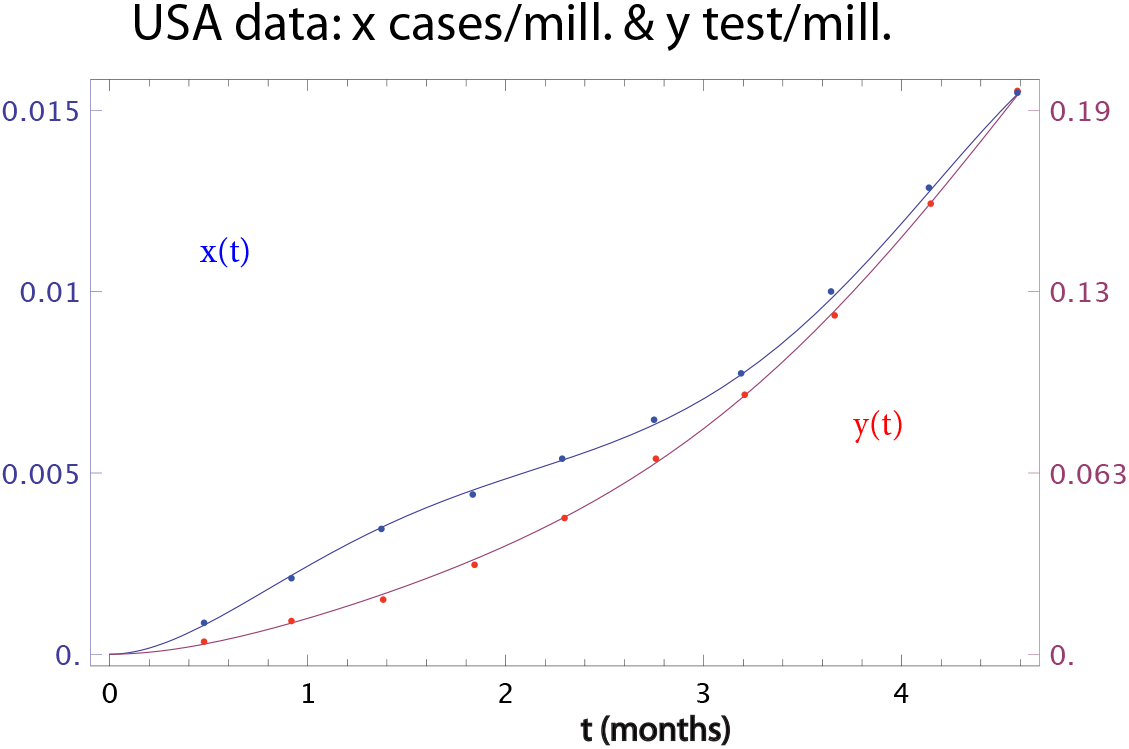

We then solve PRE for *P* with various values of *r*:

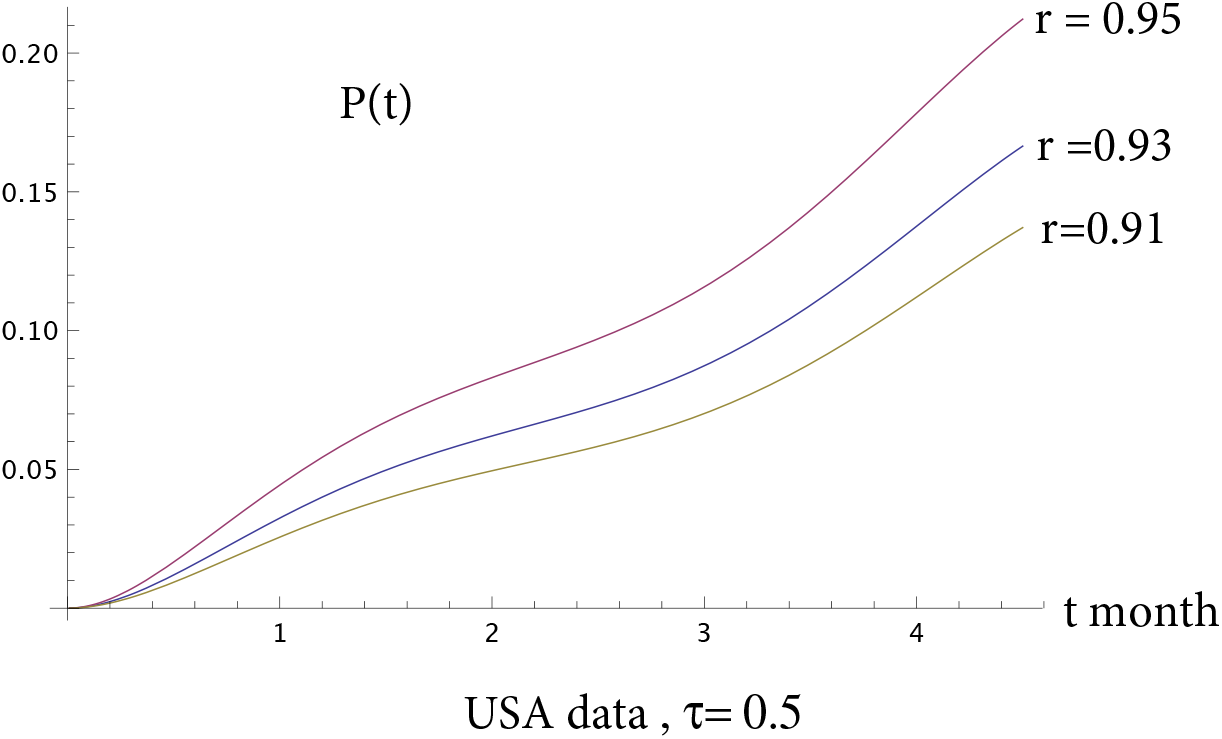

## 4 Conclusions

Our theory shows that it is possible to estimate the proportion of all who have been infected by methods easy enough to perform on any computer with *Mathematica*. The estimate is as accurate as sampling, and stable. Thus, it is not necessary to sample the whole population.

Using this method, one can make predictions on the saturation point, i.e. the proportion *σ* of susceptibles in the population. Of course, this depends also on the population profile, in particular the amount of older, sick people. Applying our formula to data from the states NY and NJ one sees that *P >* 30%, which suggests that *σ* lies in the range 35–45%, which is consistent with their *x* curves.

The formula shows proportion presently infected is

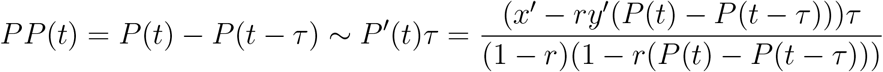

Various authorities have tried to estimate *P P*: they use the proportion of infected among those daily tested, analytically this is the function *x*′ (*t*)/*y*′ (*t*).

Comparing *x*′ (*t*)/*y*′ (*t*) with *P′ (t) τ* for USA data:

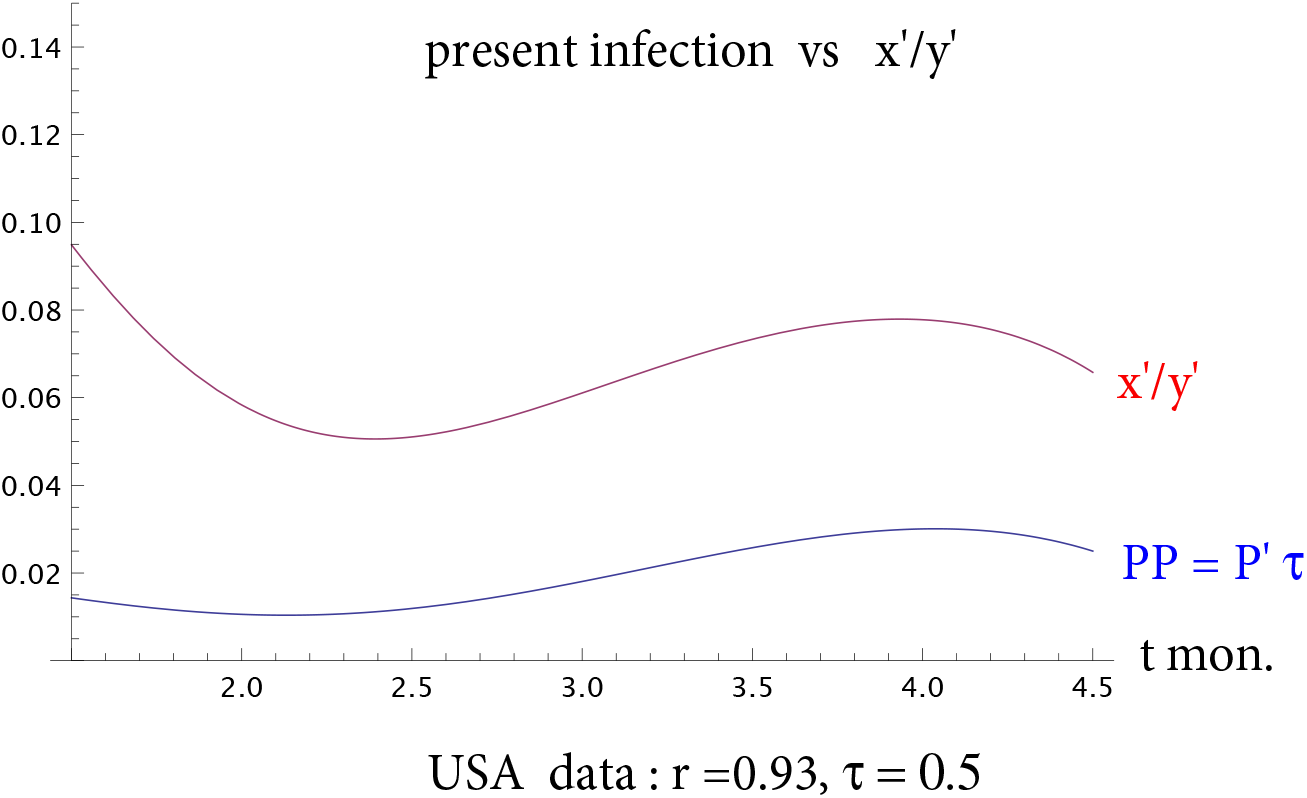

At the present time, the *x*′ (*t*)/*y*′ (*t*) estimate is out by about a factor two, but nonetheless, in the right ballpark (and erring on the safe side).

The CDC has used *x/P* to estimate *ρ*. This is accurate enough for small *t*, but for current time, our theory shows, with *τ ∼* 0.5, that the CDC method underestimates *ρ* by at least 20%, with the discrepancy growing with time.

One could also make quick estimates of *P* for various nations. Our main example is the USA, which at *P ∼* 18%, has not too far to go before reaching saturation. On the other hand, my native Australia, which had a severe lockdown, seems to have *P ∼* 1%, i.e. it has a long, long way to go.

## 5 Data and computations

There is considerable data available from CDC, Oxford University [4], and Los Alamos[6]. All computations done on an iMac using *Mathematica*.

## Data Availability

All available from author at

## Author Declarations

*I confirm all relevant ethical guidelines have been followed, and any necessary IRB and/or ethics committee approvals have been obtained.*

*The details of the IRB/oversight body that provided approval or exemption for the research described are given below:*

*All necessary patient/participant consent has been obtained and the appropriate institutional forms have been archived.*

*I understand that all clinical trials and any other prospective interventional studies must be registered with an ICMJE-approved registry, such as ClinicalTrials.gov. I confirm that any such study reported in the manuscript has been registered and the trial registration ID is provided (note: if posting a prospective study registered retrospectively, please provide a statement in the trial ID field explaining why the study was not registered in advance).*

*I have followed all appropriate research reporting guidelines and uploaded the relevant EQUATOR Network research reporting checklist(s) and other pertinent material as supplementary files, if applicable.*

## Footnotes

[2] Our first version[1] used another model, where symptomatic cases are not immediately tested. We found that even with a delay of two weeks, there was no significant difference from the present immediate response model-whose mathematics is much easier to handle.

[3] In [1] we considered the plausible idea that there was delay *∼ τ*.

